# Impact of blood pressure control level on all-cause mortality and CVD mortality in patients with hypertension and its comorbidities: A prospective cohort study in Southwest China

**DOI:** 10.1101/2024.05.30.24308250

**Authors:** Yanli Wu, Ling Li, Jie Zhou, Ji Zhang, Yiying Wang, Lisha Yu, Tao Liu

## Abstract

**Background:** Blood pressure (BP) control levels may be associated with the risk of all-cause mortality and CVD mortality. The goal of this study was to explore the association of BP control levels with the risk of all-cause mortality and CVD mortality in adults in Guizhou, China.

**Methods:** A 13-year prospective cohort study of 1,905 hypertension patients adults aged 18 years or older was conducted in Guizhou, southwest China from 2010 to 2023. Information of participants death were collected, cause of death was coded according to ICD-10 and codes I00-I99 was categorized as CVD-related deaths. Cox proportional hazard regression was used to examine the associations of BP control levels with all-cause mortality and CVD mortality.

**Results:** 134 deaths occurred during a median follow-up of 11.72 years. The multivariable cox hazard regression models showed that compared with SBP≥140 mmHg and DBP≥90 mmHg group, participants with SBP <120 mmHg and DBP <80 mmHg, they had the lowest risk of all-cause mortality (HR=0.265, 95%CI:0.111,0.630) and the lower risk of CVD death (HR=0.274, 95%CI:0.092,0.814). Associations between BP control levels with all-cause mortality and CVD mortality were suggested to be even stronger among aged ≥65 years (*P*<0.05), but ideal BP levels were not significantly associated with CVD mortality in participants with comorbidity (*P*>0.05).

**Conclusion:** Optimal BP control levels(<120/80mmHg) is effective in reducing overall mortality and cardiovascular disease (CVD) mortality, even among older adults. however, for hypertension with comorbidities, mere BP control alone may not suffice, consideration should also be given to the treatment of other coexisting conditions.

**NOVELTY AND RELEVANCE:** *What Is New?:* To our knowledge, few studies have investigated the relationship between the levels of blood pressure control in hypertensive patients and all-cause mortality in the Chinese population. Furthermore, there exists divergent viewpoints regarding the extent of blood pressure management in geriatric patients diagnosed with hypertension.In this study, we used data from a prospective cohort study in Southwest China to investigate whether the level of blood pressure control of hypertension patients was associated with all-cause mortality and cardiovascular disease (CVD) death and test whether those associations were modified by age or comorbidity factors.

*What Is Relevant?:* Using data from a prospective cohort study in Southwest China, we found that maintaining optimal blood pressure control levels (<120/80mmHg) was found to significantly reduce overall mortality and cardiovascular disease (CVD) mortality, even among elderly individuals.

*Clinical/Pathophysiological Implications?:* The association between blood pressure and all-cause mortality serves as a guide for the management and control of hypertension in patients.

---

Hypertension is a leading risk factor of cardiovascular disease and it is becoming increasingly prevalent globally, and it is also the main cause of premature death worldwide^[1]^. With the aging of the population and the change of residents’ lifestyle, the number of hypertension patients continues to increase, and has become an important public health problem**^Error! Reference source not found.^**. An estimated 1.28 billion adults aged 30-79 years worldwide have hypertension, most (two-thirds) living in low- and middle-income countries, 46% of adults with hypertension are unaware that they have the condition, and approximately 1 in 5 adults (21%) with hypertension have it under control^[1]^. It is reported, In 2021, about 1 in 4 of Chinese adults suffered from hypertension, but the awareness rate and treatment rate were <50%, and the control rate was only 15%^[2]^. The 2019 Global Burden of Disease Study (Global Burden of Disease Study 2019, GBD 2019) estimated that about 10.85 million deaths due to hypertension worldwide in 2019, accounting for 31% of all causes of death^[4]^. Therefore, the burden of death due to hypertension remains high.

The World Health Organization recommends that the blood pressure goal in patients with hypertension is less than 140/90 mmHg, and it is less than 130/80 mmHg if you have cardiovascular disease or diabetes, etc^[1]^. However, The optimal goal of blood pressure control in elderly patients with hypertension has been highly controversial, Many scholars believe that the blood pressure control target for the elderly patients with hypertension should be appropriately relaxed to <140/90∼150/90 mmHg^[5]^. Numerous studies have shown that the control rates of hypertension is low^[2],[6]^**^Error! Reference source not found.^**, and its have a large age difference^[7]^. To our knowledge, studies on hypertension and death have mostly focused on the risk of death in hypertensive patients^[9]-[11]^, few studies have investigated the relationship between the levels of blood pressure control in hypertensive patients and all-cause mortality in the Chinese population.

It is necessary to clarify this relationship to provide a scientific basis for the prevention and treatment of hypertension. In this study, we used data from a prospective cohort study in Southwest China to investigate whether the level of blood pressure control of hypertension patients was associated with all-cause mortality and cardiovascular disease (CVD) death and test whether those associations were modified by age or comorbidity factors.

## METHODS

### Study Population

The data were obtained from the Guizhou Population Health Cohort Study (GPHCS), a large population database that aimed to investigate the epidemic of chronic diseases and risk factors. Briefly, a total of 9280 adult residents from 48 townships of 12 districts were recruited into this prospective cohort using the multistage proportional stratified cluster sampling method. The eligibility criteria of subjects included those who (1)were aged 18 years or above, (2) lived in the study region and had no plan to move out, (3) completed survey questionnaire and blood sampling, and (4) signed the written informed consent form. Based on the intent-to-treat criteria, participants were followed up from the date of entry until death, loss to follow-up, time of a request for no further contact, or until the first planned completion date in 2016-2020. During the first follow-up until 2020, information were updated on the status of major chronic diseases and vital status^[12]^. participants were followed up secondly in 2023, and the information were updated about vital status, with a response rate of 99%.

In this study, we excluded 5 respondents with missing data on hypertension and 6814 participants without hypertension at baseline, and 556 participants missing data on pressure value at the first followed up. Finally, the remaining 1,905 participants were included in the analysis (Figure 1). The study was approved by the Institutional Review Board of the Guizhou Center for Disease Control and Prevention (No. S2017-02) and written informed consent was obtained from all participants. Personal sensitive information involved in the database (such as phone number, identification number, home address, etc.) has been deleted during the data analysis process in oder to protect the participants’ personal information and health data, and database was protected according to Data Security Law of the People’s Republic of China.

**Figure 1.**
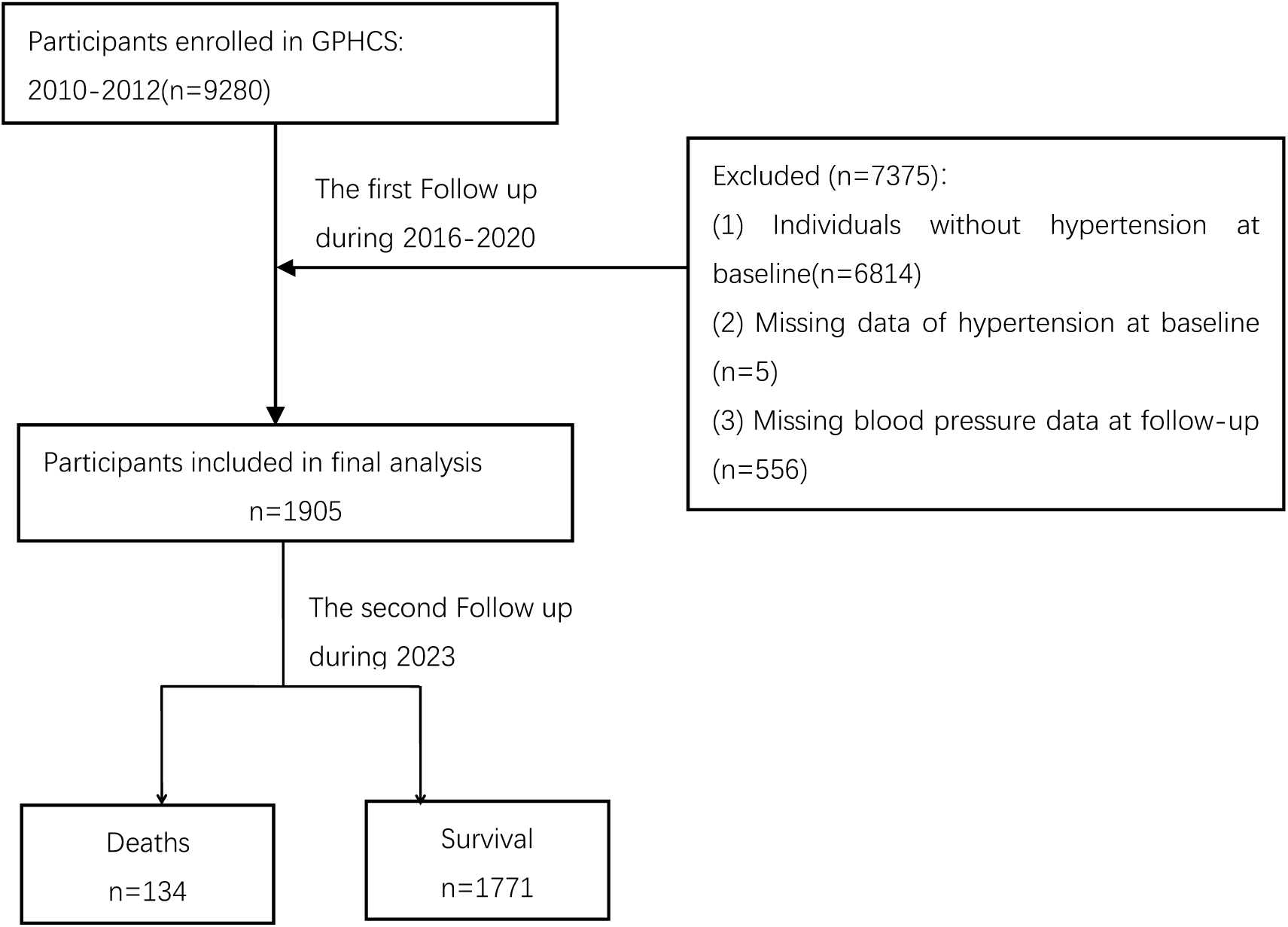
The flow chart of the study.

### Assessment of Hypertension

hypertension included those with a systolic blood pressure≥140 mmHg and/or diastolic blood pressure≥90 mmHg, or self-reported hypertension^[13]^.

### The Level of Blood Pressure Control and Grouping

In this study, the blood pressure values of the first follow-up were taken as the level of blood pressure control of participants. The individuals were split into five categories according the level of blood pressure control: ①SBP≥140 mmHg and DBP≥90 mmHg, ②SBP 120-139 mmHg and DBP 80-89 mmHg, ③SBP<120 mmHg and DBP<80 mmHg, ④SBP≥140 mmHg and DBP< 90 mmHg, ⑤SBP<140 and DBP≥90.

### Ascertainment of Death

Information of participants death were collected through cover the province’s resident cause of death monitoring system containing complete case information such as ID number, immediate cause of death and underlying cause of death or through being informed by a family member. Coded cause of death was coded according to the International Statistical Classification of Diseases and Related Health Problems, Tenth Revision (ICD-10)**^Error! Reference source not found.^**. For the present analyzes, we categorized codes I00-I99 as CVD-related deaths.

### Assessment of Covariates

Information on the covariates were collected through face-to-face questionnaires survey and physical measurements by trained health workers^[14]^. The survey included sociodemographic characteristics (age, sex, ethnicity, region, and marriage status), lifestyle habits (smoking status, alcohol use, diet, and physical activity), history of obesity, diabetes, dyslipidemia, and CVD. Physical measurements including height, body weight, and blood pressure. Never smoker was defined as those who had never smoked. Never drinker was defined as those who had never consumed alcohol. Inadequate fruit and vegetable intake was defined as fruit and vegetable intake of less than 400g/day. Physical activity was defined as having moderate or vigorous physical activity at least 10 min every time for three or more times per week. BMI was calculated by dividing the weight by height squared (kg/m^2^), obesity was defined as BMI≥28 kg/m^2[15]^. After at least 8 h of overnight fasting, a 75 g 2h oral glucose tolerance test was conducted for each participant. A venous blood sample was collected before and 2 h after glucose administration. Plasma glucose was detected using the hexokinase method within 4 h. After centrifugation, serum separated from the remaining blood samples was stored at −20℃ to detect levels of fasting plasma glucose (FPG), 2-h glucose, triglycerides (TG), total cholesterol (TC), low-density lipoprotein cholesterol (LDL-C), and high-density lipoprotein cholesterol (HDL-C) (Olympus 400 Analyzer; Beckman Coulter, Brea, CA, USA). Diabetes was defined according to according to the 1999 World Health Organization criteria^[17]^: 1) having been diagnosed with diabetes by township or community and above hospitals, 2) fasting plasma glucose (FPG)≥7.0 mmol/l (126 mg/dl), 3) 2-h glucose≥11.1 mmol/l (200 mg/dl). Dyslipidemia was defined as who met either of the following criteria^[18]^: (1) self-reported doctor diagnosis of dyslipidemia or use of lipid regulating drugs; (2) high TC: TC ≥6.22 mmol/L; (3) high TG: TG≥2.26 mmol/L; (4) low HDL-C: HDL-C <1.04 mmol/L; 5) high LDL-C: LDL-C≥4.14 mmol/L. Lastly, CVD and cancer was defined as those who self-reported having CVD or cancer.

### Statistical Analysis

The Statistical Package for the Social Sciences (version 26.0; IBM Corporation, Armonk, NY, USA) and R software (Version 4.1.2; R Foundation for Statistical Computing, Vienna, Austria) were used to perform statistical analyses. Data were described as means and SDs for continuous variables, and as frequencies and percentages for categorical variables. Baseline characteristics were compared via the analysis of variance or the Chi-square test. Person-years were used as the time variable. The person-years were calculated from the baseline survey to death or the end of follow-up. Cox proportional hazards regression was used to calculate associated hazard ratios (HRs) and 95% CIs, adjusted for baseline covariates.

There sensitivity analyses were conducted to assess the robustness of the results: ① Excluding participants with CVD in baseline, ② excluding participants who died within the first 6 years of follow-up, and ③ we adjusted for ethnicity where such data were available.

The Schoenfeld residuals were used to test the assumption of hazard proportionality in Cox regression models and found no evidence of non-proportionality (*P*≥0.05). All analyses were two-tailed and P < 0.05 was considered to indicate statistical significance.

## RESULTS

### Baseline Characteristics of Participants

During a median follow-up of 11.72 years, 1905 participants at baseline were included in the analysis. Of all subjects, the average age was 52.68±14.23 years old and more than half were men(52.13%). During the follow-up period, 134 deaths occurred during the 13-year follow-up period. According to whether deaths occurs at follow-up The baseline characteristics of participants are presented in Table 1. Compared with non-death group, death group more likely to be older, Education years ≥9 years, non-married, they also had a higher proportion of cancer and smoking and a higher values of SBP and PBG. However, the proportion of dyslipidemia and BMI were lower in death group than non-death group.

**Table 1.** The baseline characteristics of the study population by vital status.

| | Total (n=1905) | Alive (n=1771) | dead (n=134) | $\chi^2$ | P |
| --- | --- | --- | --- | --- | --- |
| Age≥45 years, % | 1354(71.08) | 1229(69.40) | 125(93.28) | 43.578 | <0.001 |
| Men, % | 993(52.13) | 916(51.72) | 77(57.46) | 1.645 | 0.200 |
| Urban, % | 602(31.60) | 554(31.28) | 48(35.82) | 1.187 | 0.276 |
| Han ethnicity, % | 1164(61.10) | 1084(61.21) | 80(59.70) | 0.119 | 0.730 |
| Education years <9 years, % | 1227(64.41) | 1112(62.79) | 115(85.82) | 28.826 | <0.001 |
| Married, % | 1544(81.05) | 1452(81.99) | 92(68.66) | 14.414 | <0.001 |
| Never drinker, % | 1244(65.30) | 1155(65.22) | 89(66.42) | 0.079 | 0.778 |
| Never smoker, % | 1294(67.93) | 1214(68.55) | 80(59.70) | 4.476 | 0.034 |
| Insufficient intake of fruits and vegetables, % | 1290(67.93) | 1203(68.16) | 87(64.93) | 0.598 | 0.439 |
| Physical activity, % | 139(7.30) | 130(7.34) | 9(6.72) | 0.072 | 0.789 |
| Dyslipidemia, % | 1281(67.24) | 1202(67.87) | 79(58.96) | 4.496 | 0.034 |
| Diabetes, % | 268(14.07) | 249(14.06) | 19(14.18) | 0.003 | 0.995 |
| CVD, % | 17(0.89) | 16(0.90) | 1(0.75) | 0.035 | 0.852 |
| Obesity, % | 259 (13.63) | 247 (13.99) | 12 (8.96) | 2.678 | 0.102 |
| Cancer, % | 5(0.26) | 3(0.17) | 2(1.49) | 8.319 | 0.004 |
| Age, years | 52.68±14.23 | 51.76±13.98 | 64.8±11.78 | -12.181 | <0.001 |
| BMI, kg/m<sup>2</sup> | 24.08±3.64 | 24.15±3.63 | 23.07±3.63 | 3.337 | 0.001 |
| DBP, mmHg | 91.44±11.8 | 91.46±11.77 | 91.23±12.3 | 0.213 | 0.831 |
| SBP, mmHg | 151.16±20.14 | 150.72±19.96 | 156.91±21.66 | -3.438 | 0.001 |
| FBG, mmol/L | 5.42±1.39 | 5.40±1.35 | 5.62±1.83 | -1.730 | 0.084 |
| PBG, mmol/L | 6.13±2.61 | 6.08±2.51 | 6.89±3.62 | -2.514 | 0.013 |
| CHOL, mmol/L | 4.98±1.34 | 4.98±1.35 | 4.98±1.21 | -0.022 | 0.983 |
| TG, mmol/Ls | 2.09±1.75 | 2.10±1.74 | 2.00±1.92 | 0.614 | 0.539 |
| HDL, mmol/L | 1.44±0.61 | 1.44±0.62 | 1.44±0.54 | -0.023 | 0.982 |
| LDL, mmol/L | 2.72±1.23 | 2.72±1.24 | 2.80±1.11 | -0.716 | 0.474 |

### Blood Pressure Control Levels and Risk of All-cause Mortality and CVD Mortality

In our study, 134 deaths occurred during the 13-year follow-up period, including 64 CVD deaths. The overall incidence density rate of all-cause mortality was 5.89 per 1000 personyears. In the last 13 years, the incidence rates of all-cause and CVD deaths mortality among participants with SBP≥140 and DBP≥90 were significantly higher than other blood pressure groups (table 2). The unadjusted (Table 2, Model 1) and sociodemographic confounders (age, sex, region, marriage, education years) and lifestyle adjusted (smoking status, alcohol use, physical activity, vegetables and fruits intake status)(Table 2, Model 2) Cox model showed that the level of the blood pressure control was associated with an decreased risk of all-cause mortality and CVD death. Compared with SBP≥140 mmHg and DBP≥90 mmHg group, after further adjusting for obesity, diabetes, CVD, cancer and dyslipidemia, participants with SBP 120-139 mmHg and DBP 80-89 mmHg had 0.523 times (HR=0.523, 95%CI:0.339,0.805) risk of all-cause mortality and had 0.395 times (HR=0.395, 95%CI:0.211,0.738) risk of CVD death. When the subjects’ blood pressure was controlled to SBP <120 mmHg and DBP <80 mmHg, they had the lowest risk of all-cause mortality (HR=0.265, 95%CI:0.111,0.630) and the lower risk of CVD death (HR=0.274, 95%CI:0.092,0.814). If the individuals’ blooed pressure values were SBP≥140 mmHg and DBP <90 mmHg, it was still able to reduce the risk of all-cause mortality and CVD death, the HRs was 0.421(95% CI:0.259, 0.686) and 0.260(95% CI:0.124, 0.544) respectively. Nevertheless, the association between blood pressure of SBP<140 mmHg and DBP≥90 mmHg and all-cause mortality was not found in this study (Table 2, Model 3,Figure 2).

**Figure 2.**
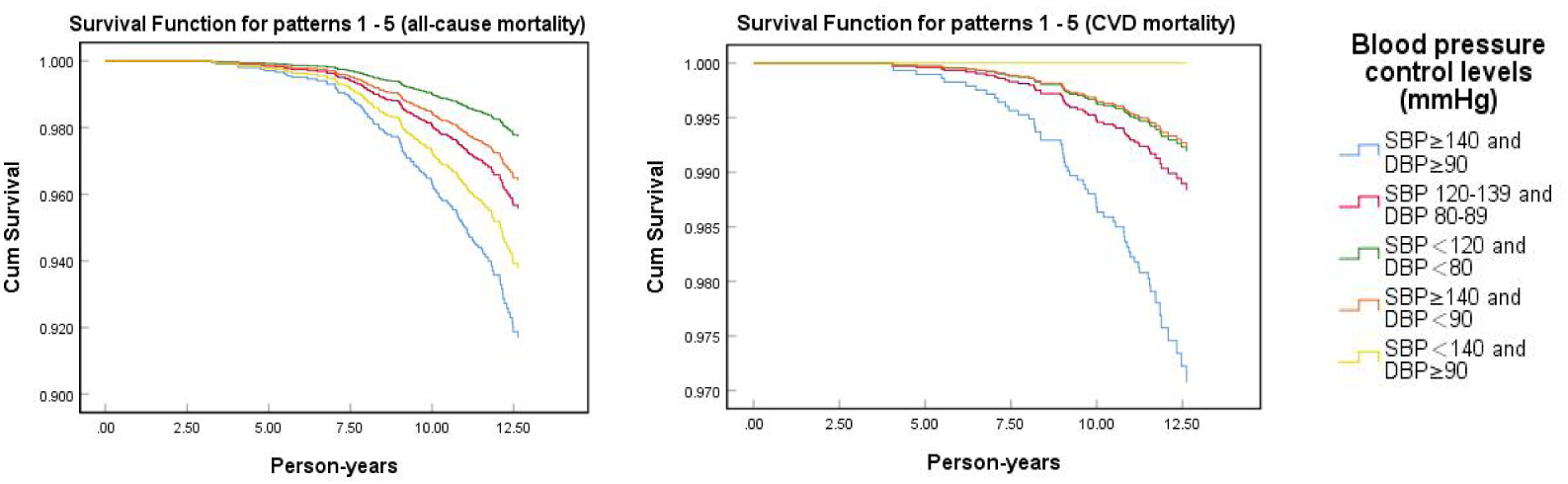
The suvival of all-cause mortality and CVD mortality at varying levels of blood pressure control. Adjusted age, sex,region, education years, marriage, smoking status, alcohol use, physical activity, obesity, CVD, cancer, diabetes, dyslipidemia.

**Table 2.** Blood pressure control levels and risk of all-cause mortality and CVD mortality.

| Blood pressure control levels(mmHg) | n | No. of case (%) | Model 1<br>*HRs* (95% CI) | Model 2<br>*HRs* (95% CI) | Model 3<br>*HRs* (95% CI) |
| --- | --- | --- | --- | --- | --- |
| all-cause mortality |  |  |  |  |  |
| SBP≥140 and DBP≥90 | 396 | 43(10.86) | 1.000 | 1.000 | 1.000 |
| SBP 120-139 and DBP 80-89 | 817 | 49(6.00) | 0.575(0.382, 0.866) | 0.548(0.362, 0.829) | 0.523(0.339, 0.805) |
| SBP<120 and DBP<80 | 222 | 7(3.15) | 0.306(0.138, 0.681) | 0.291(0.130, 0.652) | 0.265(0.111, 0.630) |
| SBP≥140 and DBP<90 | 417 | 32(7.67) | 0.702(0.445, 1.110) | 0.449(0.283, 0.712) | 0.421(0.259, 0.686) |
| SBP<140 and DBP≥90 | 53 | 3(5.66) | 0.551(0.171, 1.777) | 0.729(0.224, 2.374) | 0.741(0.225, 2.442) |
| *P* for trend |  |  | 0.015 | 0.002 | 0.001 |
| CVD mortality |  |  |  |  |  |
| SBP≥140 and DBP≥90 | 396 | 25(6.31) | 1.000 | 1.000 | 1.000 |
| SBP 120-139 and DBP 80-89 | 817 | 22(2.69) | 0.443(0.25, 0.786) | 0.415(0.232, 0.743) | 0.395(0.211, 0.738) |
| SBP<120 and DBP<80 | 222 | 5(2.25) | 0.374(0.143, 0.977) | 0.313(0.118, 0.832) | 0.274(0.092, 0.814) |
| SBP≥140 and DBP<90 | 417 | 12(2.88) | 0.453(0.228, 0.903) | 0.260(0.130, 0.521) | 0.260(0.124, 0.544) |
| SBP<140 and DBP≥90 | 53 | 0(0) | - | - | - |
| *P* for trend |  |  | 0.032 | 0.001 | 0.002 |

Note: "-" indicates that there were no deaths in this group.
Model 1: Unadjusted;
Model 2: adjusted age, sex, region, education years, marriage, smoking status, alcohol use, physical activity;
Model 3: Model 2 plus obesity, CVD, cancer, diabetes, dyslipidemia.

### The Analysis Stratified by Age Group

In the analysis stratified by age (Table 3), the HRs of all-cause mortality of the controlled blood pressure groups (SBP 120-139 mmHg and DBP 80-89 mmHg group, SBP<120 mmHg and DBP<80 mmHg group) were all lower (HR_SBP 120-139 and DBP 80-89_=0.0.367, 95%CI:0.201,0.670, HR_SBP_<_120 and DBP_<_80_=0.150, 95%CI:0.044,0.512) than uncontrolled group (SBP≥140 and DBP≥90) in participants with aged 65 years or older. The all-cause mortality risk of SBP≥140 mmHg and DBP<90 mmHg groups was also significantly lower than SBP≥140 mmHg and DBP≥90 mmHg group (HR=0.367, 95%CI:0.158,0.568) for participants with aged 65 years or older. The associations between the levels of blood pressure control and CVD death were also found only in participants with aged 65 years or older. The HRs of SBP 120-139 mmHg and DBP 80-89 mmHg group, SBP<120 mmHg and DBP<80 mmHg group and SBP≥140 mmHg and DBP<90 mmHg group were respectively 0.336(95%CI:0.151,0.748), 0.244(95%CI:0.067,0.885), 0.238(95%CI:0.099,0.572).

**Table 3.**
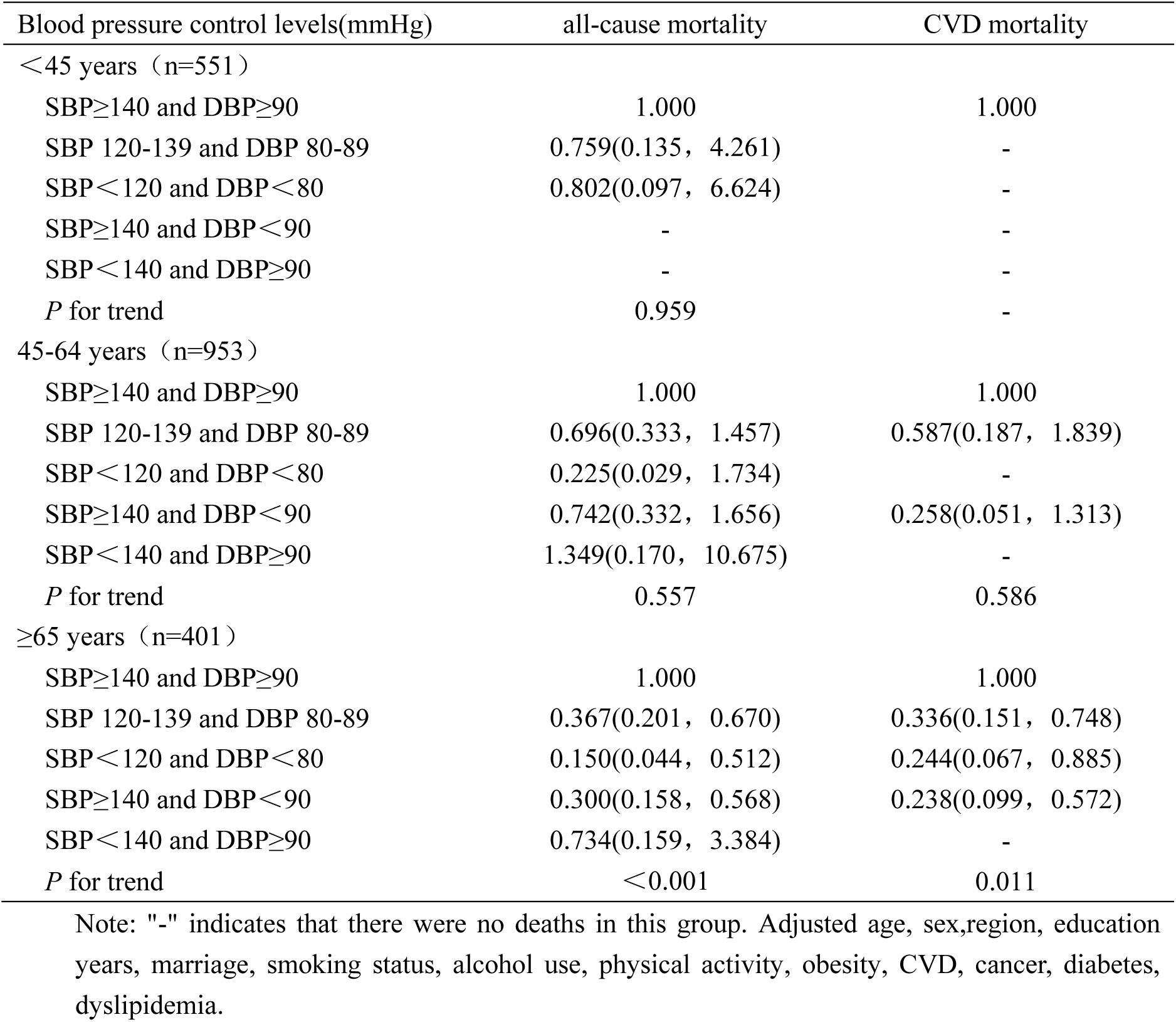
Association of blood pressure levels with all-cause mortality and cardiovascular disease death stratified by age.

### Effect of BP Control Levels on Death in Patients with Hypertension and Its Comorbidities

This study executed stratified analysis according to participants with comorbidity or not (Figure 3). a stronger association between the levels of blood pressure control and all-cause mortality and CVD mortality was found for non-comorbidity than comorbidity. The HR for all-cause mortality in non-comorbid patients was 0.302 (95% CI:0.141–0.648) in SBP120-139 mmHg and DBP 80-89 mmHg group, 0.209(95% CI:0.045, 0.975) in SBP<120 mmHg and DBP <80 mmHg group and 0.295(95% CI:0.127, 0.687) in SBP≥140 mmHg and DBP<90 mmHg group. The HR of all-cause mortality for comorbid patients was 0.310(95% CI:0.107, 0.900) in SBP<120 mmHg and DBP<80 mmHg group and 0.499(95% CI:0.272, 0.916) in SBP≥140 mmHg and DBP<90 mmHg group. In controlled blood pressure groups, Participants without comorbidity also had lower HRs for CVD mortality than those with comorbidity.

**Figure 3.**
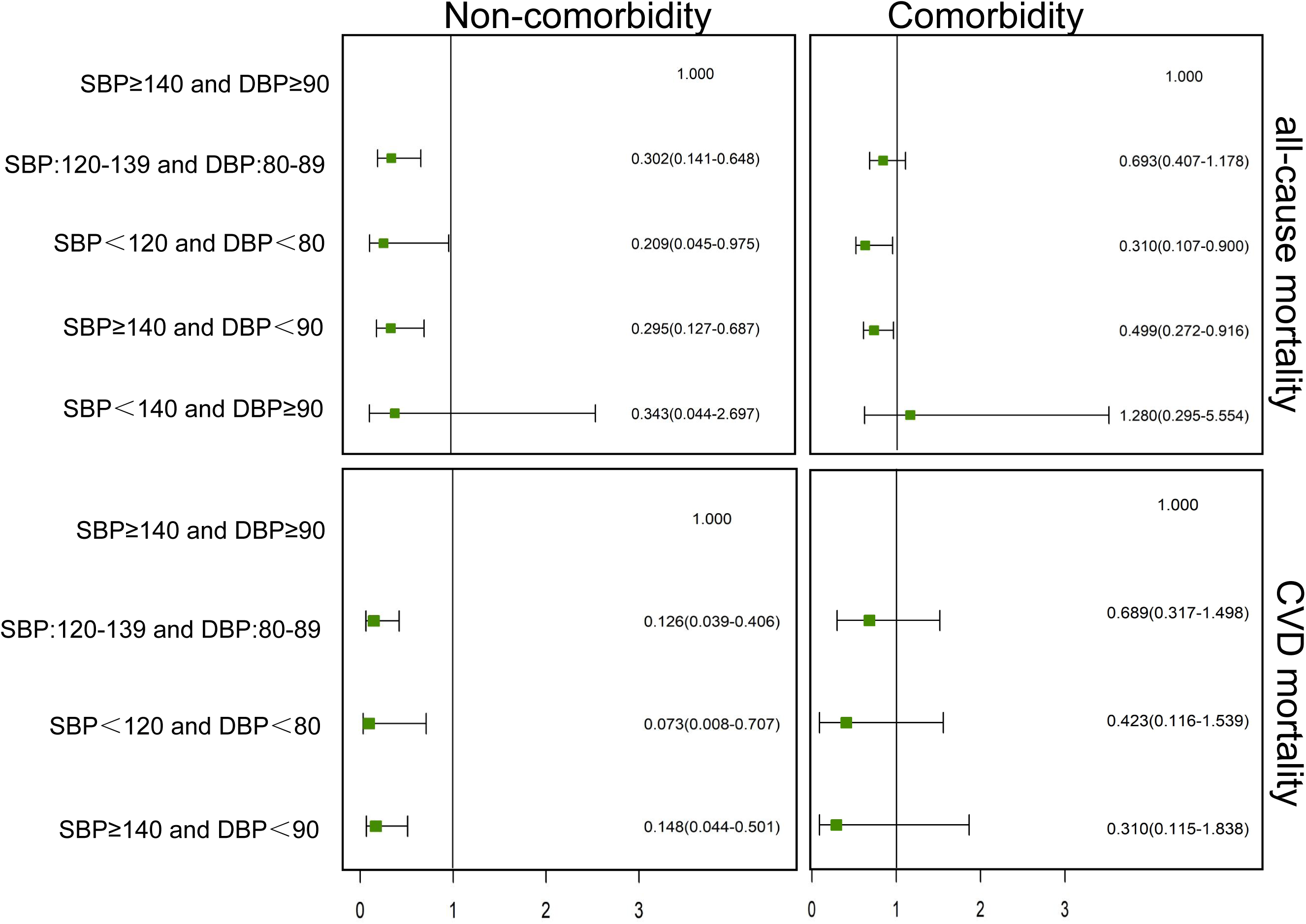
Effect of BP control levels on death in patients with hypertension and its comorbidities. Adjusted age, sex,region, education years, marriage, smoking status, alcohol use, physical activity, obesity, CVD, cancer, diabetes, dyslipidemia.

### Sensitivity Analysis

Three sensitivity analysis were conducted by excluding participants who died within the first 6 years of follow-up, excluding participants who were diagnosed CVD in baseline and adjusting for ethnicity as a categorical variable, and the results of which did not differ substantially from those of the primary analysis (Table 4).

**Table 4.**
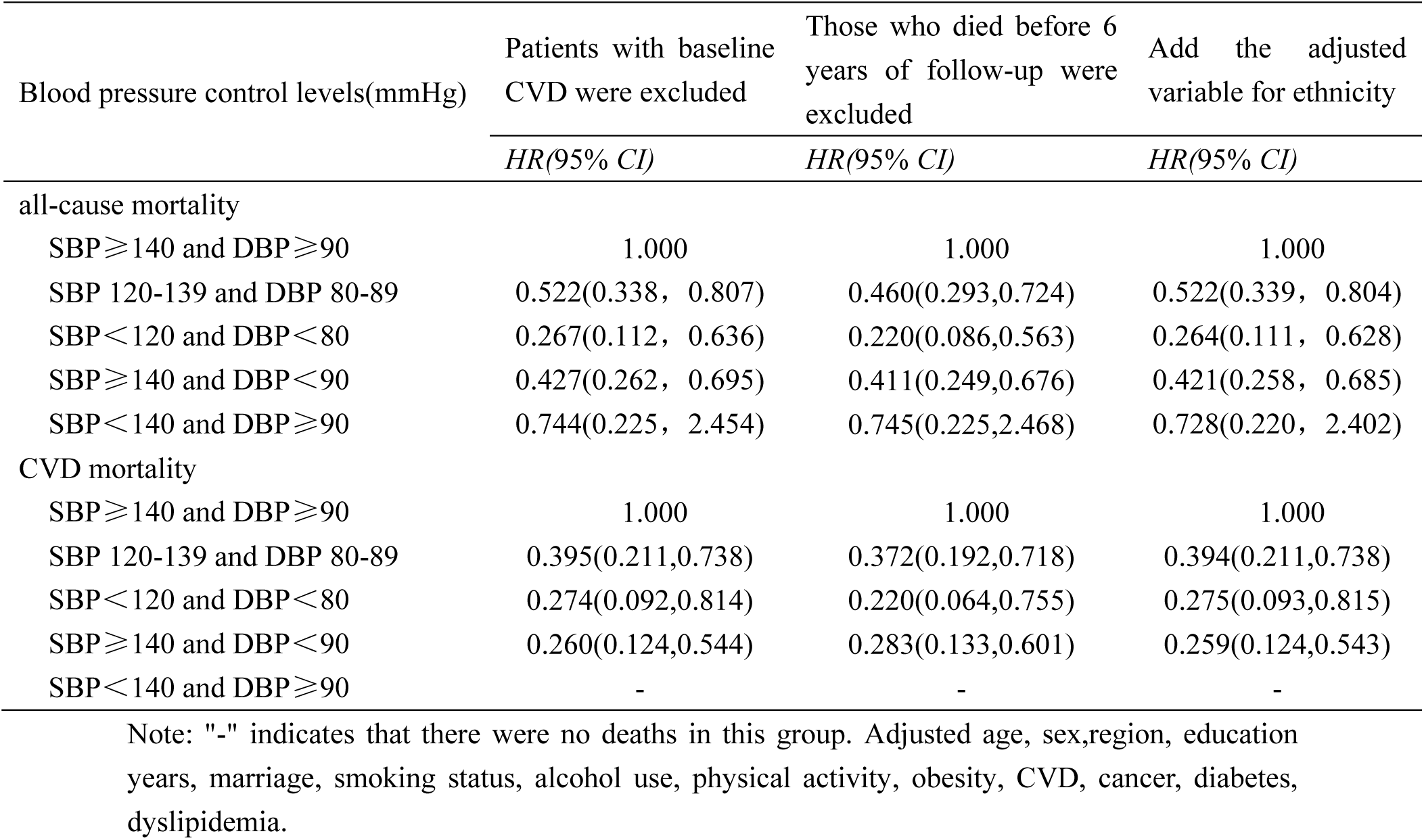
Sensitivity analysis.

| Blood pressure control levels(mmHg) | Patients with baseline<br>CVD were excluded | Those who died before 6<br>years of follow-up were<br>excluded | Add the adjusted<br>variable for ethnicity |
| --- | --- | --- | --- |
|  | *HR(95% CI)* | *HR(95% CI)* | *HR(95% CI)* |
| all-cause mortality |  |  |  |
| SBP≥140 and DBP≥90 | 1.000 | 1.000 | 1.000 |
| SBP 120-139 and DBP 80-89 | 0.522(0.338, 0.807) | 0.460(0.293,0.724) | 0.522(0.339, 0.804) |
| SBP<120 and DBP<80 | 0.267(0.112, 0.636) | 0.220(0.086,0.563) | 0.264(0.111, 0.628) |
| SBP≥140 and DBP<90 | 0.427(0.262, 0.695) | 0.411(0.249,0.676) | 0.421(0.258, 0.685) |
| SBP<140 and DBP≥90 | 0.744(0.225, 2.454) | 0.745(0.225,2.468) | 0.728(0.220, 2.402) |
| CVD mortality |  |  |  |
| SBP≥140 and DBP≥90 | 1.000 | 1.000 | 1.000 |
| SBP 120-139 and DBP 80-89 | 0.395(0.211,0.738) | 0.372(0.192,0.718) | 0.394(0.211,0.738) |
| SBP<120 and DBP<80 | 0.274(0.092,0.814) | 0.220(0.064,0.755) | 0.275(0.093,0.815) |
| SBP≥140 and DBP<90 | 0.260(0.124,0.544) | 0.283(0.133,0.601) | 0.259(0.124,0.543) |
| SBP<140 and DBP≥90 | - | - | - |

Note: "-" indicates that there were no deaths in this group. Adjusted age, sex,region, education years, marriage, smoking status, alcohol use, physical activity, obesity, CVD, cancer, diabetes, dyslipidemia.

## DISCUSSION

In this extensive prospective cohort study, there was an inverse association between the level of blood pressure control and the risk of all-cause mortality and CVD mortality. As the the level of blood pressure control decreased, the risk of all-cause mortality and CVD mortality gradually declined, particularly among individuals aged≥65 years. Compared with those in participants with SBP≥140 mmHg and DBP≥90 mmHg, the risk of all-cause mortality and CVD mortality decreased by 73.5% and 72.6%, respectively, in participants with SBP<120 mmHg and DBP<80 mmHg; in elderly individuals, these reductions were even more pronounced at 85.0% for all-cause mortality and 75.6% for CVD mortality. At the same time, This study found that controlling diastolic blood pressure had also a significant reduction in the risk of all-cause mortality and CVD death. These findings suggest that enhancing blood pressure management among hypertensive patients may serve as a preventive measure for mitigating all-cause mortality and CVD-related fatalities.

The result of this study is similar those found by Aida Hidalgo-Benites et al.**^Error! Reference source not found.^** and Juan A Divison-Garrote et al.^[19]^, which found a higher risk of all-cause mortality among individuals who have experienced uncontrolled blood pressure or controlled but not ideal level. A recent study also shows that among individuals with normal systolic BP, cumulative DBP> 80mmHg may augment the risk of future CVD events or all-cause death**^Error! Reference source not found.^**. The relationship between blood pressure level and the risk of cardiovascular and cerebrovascular disease onset, as well as mortality, exhibits a significant causal association. A prospective observational study of 61 populations worldwide (about 1 million people, 40 to 89 years old) found a continuous, independent and direct positive relationship between clinic SBP or DBP and the risk of stroke, coronary heart disease events, and cardiovascular death**^Error! Reference source not found.^**. Another study also showed that for every 10 mmHg increase of SBP, the risk of stroke and fatal myocardial infarction increased by 53% and 31% in Asian populations**^Error! Reference source not found.^**. Blood pressure level is also closely related to the occurrence of heart failure and atrial fibrillation, long-term hypertension-left ventricular hypertrophy-heart failure and hypertension-atrial fibrillation-cerebral embolism are two important chains of cardiovascular and cerebrovascular events^[23][25]^. In addition, blood pressure levels are also closely related to end-stage renal disease**^Error! Reference source not found.^**, further increasing the risk of death.

In the age-stratified analysis, it was observed that the impact of blood pressure control levels on incident all-cause mortality and cardiovascular disease (CVD) mortality was more pronounced among participants aged ≥65 years, a lower target blood pressure level (<120/80 mmHg) was associated with reduced risks of all-cause mortality and CVD death. Although some studies have shown higher BP targets in older hypertensive patients than in general hypertensive patients^[26]^**^Error!Reference source not found.^**.

According to the National Clinical Practice Guidelines on the Management of Hypertension in Primary Health Care in China (2020), it is recommended that patients aged 65 to 79 years achieve a target blood pressure of 150/90 mmHg, which may be further reduced to 140/90 mmHg if tolerated; while patients aged 80 years and older should aim for a blood pressure of 150/90 mmHg^[30]^. And the Guidelines for Managing Hypertension in the Elderly Population in China recommend maintaining blood pressure levels below 130/80 mm Hg for non-debilitating hypertensive patients aged between 65 and 79 years, provided it is well-tolerated^[30]^. However, recent subgroup analyses also showed that lower blood pressure targets (SBP<130 mmHg) were beneficial for the elderly population^[31]^. However, this study suggests that maintaining blood pressure control below 120/80 mmHg could significantly reduce mortality rates among individuals aged 65 years and older. Therefore, it should be noted that increasing age is not a sufficient condition for setting higher blood pressure goals. For elderly patients, doctors should evaluate the treatment tolerance and possible factors of adherence to treatment according to the severity of patients’ complications, and comprehensively determine the blood pressure goals of patients. This study did not find that BP control may significantly reduce the risk of death in younger hypertensive patients. This result does not mean that young hypertensive patients do not have to control their blood pressure. It is possible that young people have better vascular elasticity and the mortality caused by hypertension is not obvious in the short term, but in the long run, blood pressure control is still needed. The findings of a prospective study conducted on individuals aged 35 to 59 years old indicate that Both hypertension and high-normal blood pressure can reduce the life expectancy of middle aged people in China^[32]^.

The results of this study indicate that for hypertensive patients with comorbid conditions including diabetes, dyslipidemia, obesity, and other ailments, adopting a lower blood pressure target is advisable in order to reduce the overall risk of mortality. Nevertheless, its effectiveness in reducing cardiovascular disease (CVD) mortality seems limited. Hypertensive patients commonly manifest hyperglycemia, often in conjunction with other metabolic cardiovascular risk factors such as obesity, dyslipidemia, hepatic steatosis, proteinuria, and hyperuricemia. The aforementioned factors contribute to the promotion and exacerbation of cardiovascular risks, ultimately leading to increased mortality rates. These findings underscore the importance for hypertensive patients with comorbidities to not only effectively manage their blood pressure but also address associated conditions such as blood lipids, blood glucose levels, BMI, among others.

The strengths of this study included the well-characterized prospective design and the longer follow-up period with a relatively low loss to follow-up rate. To our knowledge, so far, no other study has been carried out to evaluate the associations of blood pressure control levels with death risk in China. Nevertheless, several limitations should also be acknowledged. Firstly, The control level of blood pressure in this study was established based on a single visit’s blood pressure measurements, which might lead to a misclassification of the blood pressure control levels. Second, some variables, such as COPD, chronic kidney disease, were not considered due to the data restrictions. In addition, data is inevitably missing in this large-scale longitudinal representative study, and this study was conducted only in Guizhou Province, so the interpretation of the results needs to be cautious.

## PERSPECTIVES

Our findings suggest that optimal blood pressure control is effective in reducing overall mortality and cardiovascular disease (CVD) mortality, even among older adults. Simultaneously, emphasis should be placed on the management of diastolic blood pressure. Furthermore, for hypertensive patients with comorbidities, mere blood pressure control alone may not suffice; consideration should also be given to the treatment of other coexisting conditions.

## Affiliations

Guizhou Center for Disease Control and Prevention, Guiyang, China(Yanli Wu, Ling Li, Jie Zhou, Ji Zhang, Yiying Wang, Lisha Yu, Tao Liu)

## Data Availability

Application for datasets generated during and/or analysed during the current study may be considered by the corresponding author on reasonable request.

## Acknowledgements

The authors thank the participants and all who were involved in the cohort of the natural population in Guizhou province.

## Author contributions

Tao Liu conceived the study, Yanli Wu performed the statistical analysis and drafted the manuscript; Tao Liu, Ling Li, Jie Zhou, Yiying Wang, Lisha Yu, Ji Zhang critically revised the manuscript; and all authors read and approved the final version of the manuscript.

## Sources of Funding

This work was supported by Guizhou Province Science and Technology Support Program [Grant number: Qiankehe (2018)2819] and Provincial Key Construction Discipline Project of Guizhou Provincial Health Commission.

## Disclosures

None.

## Nonstandard Abbreviations and Acronyms

CVD: cardiovascular disease
BP: blood pressure
DBP: diastolic blood pressure
SBP: systolic blood pressure
BMI: body mass index
FPG: fasting plasma glucose
TG: triglycerides
TC: total cholesterol
LDL-C: low-density lipoprotein cholesterol
HDL-C: high-density lipoprotein cholesterol

## Notes

### Competing Interest Statement

The authors have declared no competing interest.

### Author Declarations

The study was approved by the Institutional Review Board of the Guizhou Center for Disease Control and Prevention (No. S2017-02) and written informed consent was obtained from all participants.

